# “It’s a moving target”: Experiences of pacing to reduce symptom exacerbation among adults living with Long COVID – Results from an international community-engaged qualitative research study

**DOI:** 10.1101/2024.12.11.24318864

**Authors:** Kiera McDuff, Darren A. Brown, Natalie St. Clair-Sullivan, Soo Chan Carusone, Kristine M. Erlandson, Lisa Avery, Ciaran Bannan, Colm Bergin, Angela M. Cheung, Richard Harding, Mary Kelly, Jessica M. Martin, Lisa McCorkell, Sarah O’Connell, Imelda O’Donovan, Margaret O’Hara, Niamh Roche, Ruth Stokes, Catherine Thomson, Liam Townsend, Jaimie H. Vera, Hannah Wei, Patricia Solomon, Kelly K. O’Brien

## Abstract

**Introduction:** Long COVID is a multisystem condition that negatively impacts daily function. Pacing is a self-management strategy to mitigate symptoms. Our aim was to describe experiences of pacing from the perspectives of adults living with Long COVID.

**Methods:** We conducted a community-engaged qualitative descriptive study involving one-on-one online interviews with adults living with Long COVID from Canada, Ireland, United Kingdom, and United States to explore experiences of disability. We asked participants about strategies they used to deal with health challenges living with Long COVID. Interviews were audio recorded and transcribed verbatim. We analyzed data using group-based content analytical techniques.

**Results:** Among the 40 participants living with Long COVID, the majority were women (n=25; 63%), white (n=29;73%) and heterosexual (n=30;75%). The median age of participants was 39 years (25th, 75th percentile: 32, 49). Most participants (n=37;93%) used pacing to mitigate or prevent symptoms. Participant described experiences of pacing across five main areas: 1) using pacing as a living strategy (pacing to mitigate multidimensional health challenges; applying pacing to many types of activities; process of pacing experienced as a moving target; pacing experienced as a helpful strategy, but not a cure for Long COVID); 2) learning how to pace (acquiring knowledge about pacing; developing strategies and skills to support pacing); 3) encountering challenges with pacing (learning how to pace; experiencing inequitable access to pacing; experiencing stigma and judgement; undergoing psychological and emotional adjustment from beliefs of ‘fighting’ or ‘pushing through’ to balancing rest with activity; making sacrifices; and encountering unexpected obstacles); 4) experiencing consequences of not pacing; and 5) conceptualising and describing pacing using analogies or metaphors.

**Discussion:** Pacing is a challenging and complex strategy used to mitigate symptoms of Long COVID. Healthcare providers should work collaboratively with patients to further refine and implement this strategy, when appropriate.

## INTRODUCTION

Long COVID, also known as Post-COVID Condition (PCC), is a prevalent and disabling chronic multisystem condition that impacts the health of millions of people (1). The National Academies of Sciences, Engineering, and Medicine (NASEM) defines Long COVID as “an infection-associated chronic condition (IACC) that occurs after a SARS-CoV-2 infection and is present for at least 3 months as a continuous, relapsing and remitting, or progressive disease state that affects one or more organ symptoms” (2). Estimates of the prevalence of Long COVID among people previously infected with SARS-CoV-2 range between 10-35% (2–4). The most common symptoms of Long COVID include extreme exhaustion or fatigue, post-exertional symptom exacerbation (PESE) or post-exertional malaise (PEM), shortness of breath, cognitive impairment (such as memory impairment, difficulty problem solving), and dizziness on sitting or standing (such as orthostatic intolerance) (5). Over 200 symptoms of Long COVID have been identified (6). The majority of people living with Long COVID report that their symptoms negatively impact their daily function and social participation (6, 7).

The symptoms and impairments of Long COVID and their impact on daily function represent disability, broadly defined as any physical, cognitive, mental and emotional health challenges, activity limitations, uncertainty about the future, and challenges to social inclusion (including return to work, family life, relationships, and ability to care for others) (8–11). For many individuals living with Long COVID, their health challenges may fluctuate over time, characteristic of episodic disability (8, 11–13). For example, PESE or PEM, the worsening of symptoms after exertion, are common symptoms of Long COVID (14, 15).

Fluctuations in disability may be unpredictable for people living with Long COVID, making it difficult to plan for the future and maintain or return to employment (8, 12, 13). Uncertainty is a dimension of disability among people living with Long COVID which may exacerbate symptoms (11). Long COVID poses substantial individual health, workforce, and economic burdens (16).

No approved treatment or cure for Long COVID exists (17). In the absence of effective treatments or cures, people living with Long COVID have adopted strategies to help manage and mitigate some of the associated symptoms and disability (18, 19). In the context of episodic disability, the behaviours, attitudes, and beliefs that patients adopt to manage their disability are referred to as living strategies (20). These living strategies include pharmacological treatments (e.g., over-the-counter medications), dietary alterations (e.g., supplements, eating low-histamine foods), and energy management (8, 21, 22). Often, people living with Long COVID adopt living strategies through their own trial and error, and may be based on advice from others living with disability (18, 19, 23).

Pacing is a living strategy that people living with Long COVID can adopt to manage their energy and navigate everyday life (18). Pacing is an approach that balances rest and activities in daily life, to manage symptoms such as fatigue and PESE or PEM (18, 24). Pacing has been utilised by people living with a range of health conditions, such as myalgic encephalomyelitis/chronic fatigue syndrome (ME/CFS), cancer, and arthritis (25). There are multiple potential goals and types of pacing within the context of energy management or conservation (24). For example, quota-contingent pacing is used to gradually increase activities, while symptom-contingent pacing guides activities based on perceived symptom levels to avoid symptom-exacerbation (26, 27). In the context of rehabilitation, it is important to consider and select the most appropriate types of pacing and energy management strategies for the patient population. For example, pacing strategies with the goal to increase activity (such as quota-contingent pacing) may not be safe or appropriate for people with Long COVID (24, 27, 28). Despite existing clinical guidelines, there is significant variability in the implementation of pacing as a living strategy among people living with Long COVID (24, 29).

There is a clinical need to understand pacing as a living strategy for people living with Long COVID, and other energy-limiting chronic conditions. It is important to understand the experiences of pacing and strategies used in the context of Long COVID to help inform clinical practice and research to mitigate disability. Our aim was to describe the experiences of pacing from the perspectives of adults living with Long COVID.

## MATERIALS & METHODS

### Study Design

We analyzed interview data from interviews collected as part of the Long COVID and Episodic Disability study, a community-engaged qualitative descriptive study involving one-on-one online interviews with adults living with Long COVID (8). The primary aim of the parent study was to describe experiences of episodic disability among adults living with Long COVID (8). Detailed methods for the Long COVID and Episodic Disability Study have been previously published (9). The study was approved by the University of Toronto Health Sciences Research Ethics Board (protocol #41749). For this study, we specifically focused our analysis on interview data pertaining to the experiences of pacing.

### Patient and Public Involvement

The Long COVID and Episodic Disability Study is a community-engaged research study involving over 25 persons living with Long COVID, researchers, and clinicians (categories are not mutually exclusive). People with lived experiences with Long COVID were involved in all phases throughout the study including conceptualisation, acquisition of funding and proposal development, recruitment and data collection, analysis and interpretation, and knowledge mobilization. During development of the study proposal, patient advocates from organisations in Canada, Ireland, UK, and US, who were linked to larger networks of people living with Long COVID, were purposefully invited to join the study team. Long COVID community networks and organisations, represented by persons living with Long COVID, were involved in all stages of the research, including: COVID Long Haulers Support Group Canada, Long COVID Ireland, Long COVID Physio, Long Covid Support, and the Patient-Led Research Collaborative (30–34). Team members with lived experiences were provided yearly remuneration for their time and expertise dedicated to the study, either as an individual, or to the community organization which they represented on the study.

### Participants and Recruitment

Participants included adults (≥18 years old) who self-identified as living with Long COVID from Canada, Ireland, United Kingdom (UK) and United States (US), defined as signs and symptoms that develop during or following an infection consistent with COVID which continue for 12 weeks or more and are not explained by an alternative diagnosis (35). Community collaborators recruited participants between December 7, 2021 and May 2, 2022 through the five online community support groups and organisations for Long COVID represented on the study team in Canada, Ireland, UK, and US (30–34). Participants in the Long COVID and Episodic Disability study were recruited to represent adults living with Long COVID across the four countries, with diversity in gender, age, and sexual orientation. All participants provided informed written or verbal consent. Participants were offered a CAN$30/€20/£15/US$20 electronic gift card for their participation in the study.

### Data Collection

Online semi-structured interviews were conducted by four members of the research team (KKO, KM, NS, BT) via Zoom between December 2021 and May 2022. We used the Episodic Disability Framework to develop the interview guide which included the following areas of inquiry: dimensions of disability; contextual factors, including living strategies such as pacing; and triggers of disability (36, 37). During the interview, participants were asked, “Did you adopt any other living or coping strategies to help you deal with the health challenges and ups and downs of disability living with Long COVID?” Following the interview, we administered a web-based questionnaire to collect demographic information (including sex, gender, race, sexuality, employment information, and Long COVID symptoms or characteristics). The interviewer followed up with each participant the day after the interview to check in on participants’ wellbeing following the interview and to provide suggestions for support services if appropriate.

### Data Analysis

We analyzed interview data from the Long COVID and Episodic Disability study that specifically pertained to the construct of pacing. We used a group-based analysis with conventional content analytical techniques (38) with portions of the interview data specific to experiences of pacing. Interview data were analysed collectively as one sample (not by country). We developed a coding framework informed by key words related to pacing (e.g. pacing, energy conservation, rest) through literature review and consultation with team members with lived experiences with Long COVID by one author (KM) while allowing for additional codes to emerge from the data. All transcripts were coded line-by-line using the coding framework. A preliminary coding summary was produced which summarised key concepts, impressions, and reflections from the data. Three team members (DAB, KKO, and KM) met to discuss and refine the preliminary coding summary. Ten team members, including five with lived experiences with Long COVID (AC, SCC, KE, SG, MK, KM, LM, MO, CT, and LT) met to review the coding summary and provide reflections, validation, and interpretation of the findings. We used NVivo software to facilitate data management (39).

## RESULTS

Forty adults living with Long COVID (10 per country) participated in interviews between December 2021 and May 2022. Participants were mostly female (n=29, 73%), white (n=29, 73%), heterosexual (n=30, 75%), with a median (25^th^, 75^th^ percentile) age of 39 years (32, 49). Thirty-three participants (83%) were living with Long COVID for a year or longer, and 34 (93%) had experienced at least one relapse in their symptoms. Of the 40 participants, 37 (93%) spoke about pacing or concepts related to pacing. Some participants explicitly named pacing (n=30;75%), while others referred to learning to incorporate more balance between rest and activity in their daily lives. See Table 1 for more details on participant characteristics.

**Table 1:** Characteristics of participants (n=40)

| Characteristics of Participants (n=40) |  | N (%) |
| --- | --- | --- |
| **Sex** |  |  |
| Female |  | 29 (73%) |
| Male |  | 11 (28%) |

| **Characteristics of Participants (n=40)** |  | **N (%)** |
| --- | --- | --- |
| **Gender** |  |  |
| Woman |  | 25 (63%) |
| Man |  | 11 (28%) |
| Non-Binary |  | 2 (5%) |
| Other: 'gender non-conforming' |  | 2 (5%) |
| **Race** |  |  |
| White |  | 29 (73%) |
| Asian, South Asian or South East Asian |  | 4 (11%) |
| Black, Caribbean, African or African American |  | 3 (8%) |
| Mixed |  | 3 (8%) |
| First Nations, Native American, or Indigenous |  | 1 (3%) |
| **Sexual Orientation / Sexuality** |  |  |
| Straight or Heterosexual |  | 30 (75%) |
| Pansexual or Queer |  | 4 (10%) |
| Gay or Lesbian |  | 3 (8%) |
| Bisexual |  | 3 (8%) |
| **Relationship Status** |  |  |
| Single (never married; never registered in civil partnership) |  | 19 (48%) |
| Married |  | 17 (43%) |
| Separated but still legally married, or divorced |  | 3 (8%) |
| In a registered civil partnership |  | 1 (3%) |
| **Highest level of education** |  |  |
| Completed university or college |  | 19 (48%) |
| Post-graduate education |  | 17 (43%) |
| Completed trade or technical training |  | 3 (8%) |
| Completed secondary / high school |  | 1 (3%) |
| **Changed Employment Since Living with Long COVID** |  |  |
| Yes (unable to work / on leave of absence due to Long COVID) |  | 20 (50%) |
| Yes (lost job or now unemployed due to Long COVID) |  | 6 (15%) |
| Yes (work reduced hours due to Long COVID) |  | 5 (13%) |
| Yes (changed employment role or job due to Long COVID) |  | 4 (10%) |
| No – Unchanged employment status |  | 5 (13%) |
| **Receives help or support because of long-term physical or mental health conditions or illnesses related to Long COVID** |  | 11 (28%) |

### Experiences of Pacing

Participants described experiences of pacing across five main areas: 1) using pacing as a living strategy; 2) learning how to pace; 3) encountering challenges with pacing, 4) experiencing consequences of not pacing, and 5) conceptualising and describing pacing using analogies and metaphors. The following sections describe each area in detail, with supporting quotations from the interviews referenced by participant number and country. See Figure 1.

**Figure 1:**
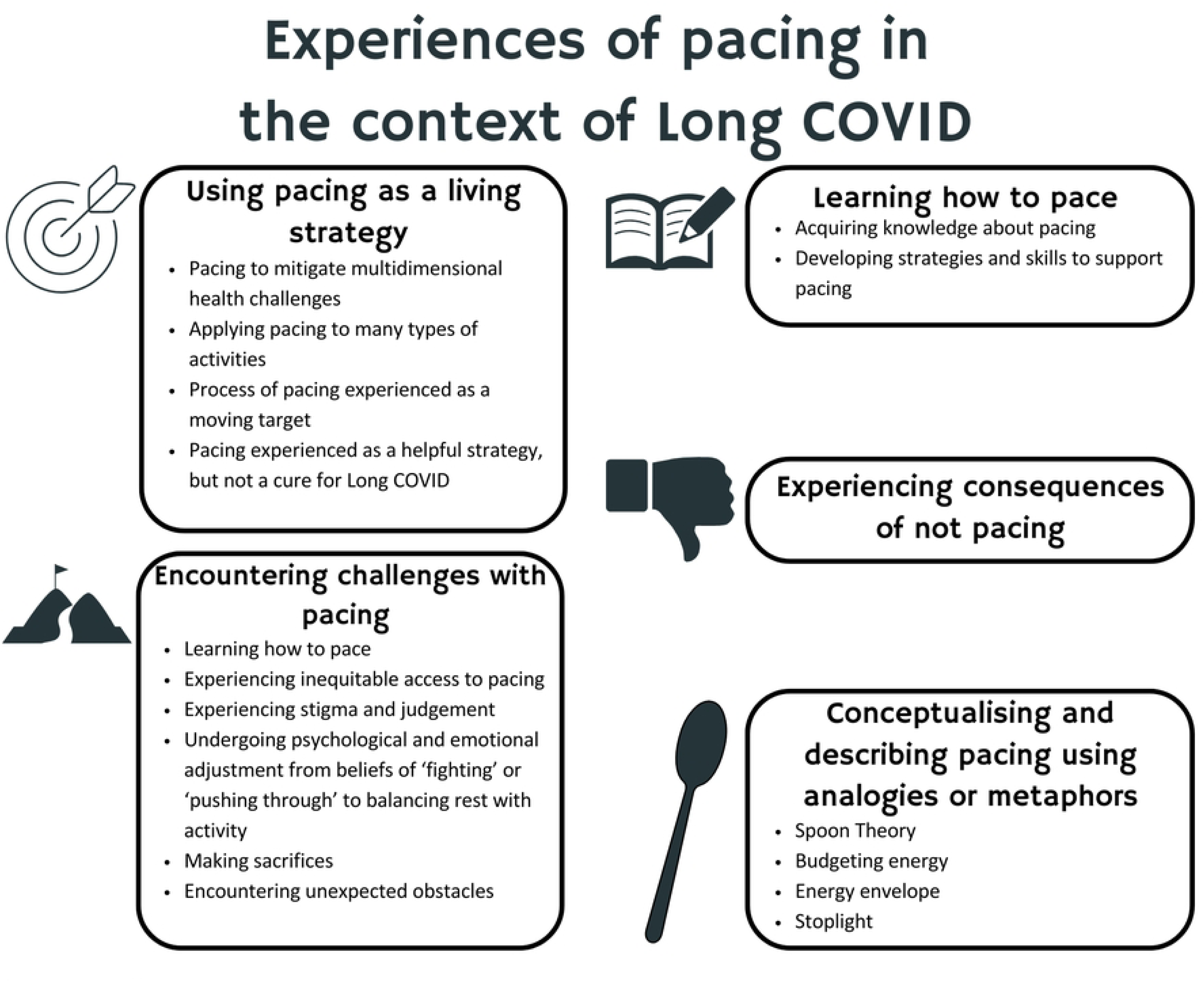
Experiences of Pacing

### 1) Using pacing as a living strategy

Participants described pacing as a living strategy to prevent, manage, or mitigate multidimensional health challenges (physical, mental, emotional, cognitive, and social) associated with Long COVID, including PESE and PEM. Pacing involved balancing activity and rest. Pacing helped participants reduce symptom severity or exacerbation, and crashes or relapses. Consistent pacing was required to maintain a lower presence and severity of symptoms. We describe further details of pacing as living strategy below.

### Pacing to mitigate multidimensional health challenges

Participants described different dimensions and factors to be considered when pacing (including emotional, cognitive, and physical exertion). Participants described pacing or limiting social interactions, such as *“visits or my phone calls”* (P29, Canada); cognitive tasks, like *“listening to a podcast”* (P26, US); as well as physical tasks (e.g. housekeeping and meal preparation).

Many participants explained that they only became aware of the energy required to complete physical, cognitive, social or emotional tasks after they developed Long COVID. One participant explained that it was easy for them to understand how they could pace physical tasks; however, realising that cognitive tasks could also be fatiguing was “*like a revelation”* (P7, Ireland). Effective pacing required finding a balance between different types of exertion. *“It’s like I have to pick one. Either I’m going to go through the mail or I’m going to do chores but I can’t do both.*” (P28, US).

### Applying pacing to many types of activities

Participants described pacing many types of activities. For participants experiencing severe impairment, the most essential activities (basic activities of daily living) had to be paced. As one participant reported: *“For a while there it was even pacing how often I go to the loo just because of that extra few steps and that little bit more energy. It’s just simple things like that you just take for granted.”* (P5, UK). Other participants were pacing day-to-day activities (such as going to appointments or managing paperwork). For example, *“If I drive somewhere, even if it’s a short distance like a 15- or 20-minute drive, I’ll have to pull in the car, park and sleep or nod off for a while before I have the energy to go the next bit.”* (P36, Ireland).

Those who were working at the time of their interview described applying pacing and energy conservation strategies in the workplace. They found they were more successful with slower and more flexible schedules so that they could spread out difficult and energy-intensive tasks. For example:

*“If I had a full day [of work] with 14 assessments in it, then it might be a different kettle of fish… If I have a day then where there’s not too much happening, that’s okay and I’m able to gain kind of energy for the next few days… I think I am pacing quite well.”* – P8, Ireland

Participants described that they had to re-learn their energy capacity when it came to the tasks involved in their jobs (e.g., more cognitively demanding vs. more passive), and pace their work days accordingly. This participant incorporated pacing to manage meetings at work:

*“…because I get shortness of breath while I’m talking in meetings, I like have someone watch my dog because if I have to like take her on walks, I can’t. I won’t be able to like do my meetings. I may ask other people to facilitate more of my meetings because I can’t talk and I’ll get shortness of breath… I have to make meetings shorter so I can like limit the air and cognitive… I’ll space meetings out as much as I can so that I can recover and… so I don’t lose as much of my cognitive functioning.”* – P35, US

Social interactions, such as visits with friends or family, and going to a coffee shop, and leisure activities also required pacing. One participant explained they were surprised that colouring could trigger an episode of PESE, likely “*related to having to think about choosing the colours and things like that… what colour do I choose and where do I paint it?”* (P30, US). They were excited to have gradually built up to 20-30 minutes of colouring at a time. Another participant described resting for two days in order to participate in a drumming class without triggering a crash. Ultimately descriptions of pacing shared by participants spanned all types of activities involving physical, emotional, or cognitive exertion.

### Process of pacing experienced as a moving target

Despite the benefits, the process of pacing was experienced as *“a moving target”* (P24, US); trial and error were required to achieve a successful balance of rest and activity, and to maintain this balance as presence and severity of symptoms changed over time. Pacing did not involve a linear or predictable progression toward less symptom burden. As this participant described, “*It’s not like I can just like say okay I rested yesterday, so I’ll be fine today… It’s just that I’d have a better chance of being able to have this chat with you.”* (P9, Ireland). Despite ongoing pacing, some participants described they had reached *“a plateau”* (P24, US) in terms of how much symptom improvement they experienced. They were facing the same energy limitations but had learned how to achieve some function within their limits.

*“The couple of hours I have now a day where I’m functioning like now, I’m functioning at a better level than I was six months ago or a year ago. But… I’m still very limited in those couple of hours a day… I’ve understood more as time’s gone on about pacing… I would say it’s a smoother ride now but that’s not because my symptoms are getting better. It’s because I’m understanding more about my limitations.”* – P19, UK

### Pacing experienced as a helpful living strategy, but not a cure for Long COVID

Rather than eliminate all symptom exacerbation and crashes associated with Long COVID, pacing was experienced as helpful for reducing the frequency or severity of crashes. One participant described that their crashes tended to be shorter “*as I’ve gotten better at managing my energy like envelope”* and *“as long as I go to bed and rest and don’t do anything”* once they begin to ‘hit their limit’ of activity (P24, US).

Participants were clear that reducing their symptoms and crashes was helpful, but did not result in recovery from Long COVID. As this participant explained:

*“People are changing their life and pacing so that their symptoms are lower. You know that’s getting better, not like spontaneously healing [from Long COVID]… I think that there’s something going on inside of us that’s causing ongoing damage and I just really feel like until that stops, you know healing [from Long COVID], that’s not going to be successful.”* – P34, US

### 2) Learning how to pace

Participants described experiences with learning how to pace. They discussed sources of information about pacing, and their successes, failures, and challenges with learning how to pace.

### Acquiring knowledge about pacing

Many participants learned about pacing through resources in online community support groups and their own trial and error. As one participant explained, *“it’s just been my own research that has helped me to find things that help”* (P8, Ireland). This involved reading, watching videos or webinars online, and staying current on research. Many participants recognised that pacing was how “*people have been doing it for years with [ME/CFS]”* (P10, Ireland). Participants sought out knowledge about pacing from community support groups or individuals online with other energy-limiting chronic conditions such as ME/CFS or Lupus.

Self-directed learning was often undertaken by participants without formal support from healthcare professionals. For some participants, they found online resources on their own before receiving any guidance or support from a health care provider. One person described that their doctor, “…*didn’t give me any advice. It was just, ‘You’ll get better with time’. Well, I’m still not better. So, I have had to learn and educate myself.”* (P34, US). They went on to say, “*When I finally got to the good Long COVID clinic and they wanted me to see someone for physical therapy, you know he was talking about pacing. But at that point I already know. I’ve done the self-education*.” (P34, US). They said the resources on ME Action’s website (40), “…*really, I think saved me from getting a lot worse”* (P34, US).

Complex and energy-limiting chronic condition communities and support groups were described as bridging the gap in formal healthcare supports for pacing and other living strategies. As the Long COVID community began to develop, specific support groups for Long COVID emerged and shared resources on pacing.

*“My best friend has lupus… So, I found out about Spoon Theory through my friend and that sort of became part of our like lexicon of conversation, just in me checking in with her. Then you know just finding that in the Long COVID community and the dysautonomia communities and then ME/CFS communities.”* – P40, US

Although many participants lacked access to healthcare providers who were knowledgeable about pacing, some learned about pacing through specialty Long COVID clinics, fatigue-management programs, or specialised providers. However, some described challenges finding specialised and knowledgeable providers, and wait times were long.

*“I’ve been to two Long COVID clinics, one a horrible experience where the doctor at the Long COVID clinic told me that my physical pain was due to anxiety when I continually told him I wasn’t experiencing any mental health issues. The second Long COVID clinic is fine. So, I’m still seeing them. Then yeah, I’m seeing now like a chronic fatigue syndrome and Long COVID like expert. The most useful advice that the people who actually know what they’re talking about have is really like just pacing.”* – P34, US

Types of health providers mentioned as providing helpful information and support for pacing included occupational therapists and physiotherapists. These providers offered individualised support and guidance on pacing.

*“I started the therapy and they tried to like space things out because they know it’s taxing on people. So that’s why they tend to do like one thing at a time. But I was also… I started working with the [occupational therapist] somewhere in there with therapy sessions as well because I needed some help around pacing. I expressed that a COVID education group wasn’t enough for me… Like the reason why I wanted to go to Integrated Chronic Care Services was because I needed help with pacing because I couldn’t figure it out for myself.”* – P38, Canada

### Developing strategies and skills to support pacing

Participants described strategies and skills that they developed to support their success with pacing. For most participants, multiple strategies or skills were implemented together to help manage symptoms living with Long COVID. For example, “*Definitely planning, pacing, resting, being mindful of what I do in my day [have helped me manage Long COVID]*” (P20, UK).

Effective pacing required high-level planning and prioritisation skills. Participants carefully identified activities that were most important and would break their weeks and days down (‘chunking’) to accomplish those tasks. The remainder of their time was dedicated to rest, which was identified as a priority and itself a meaningful activity. For example, *“If I want to see my friends… I have to plan kind of a rest day, or days”* (P32, UK). Basic activities of daily living, like meal preparation, also had to be prioritised and planned:

“*If I knew that I was going to be downstairs for most of the day… I take what I need from upstairs. So, I’ve got everything laid out for downstairs. I think ahead about what I’m cooking or you know I’d cook with little stools so I’m sitting down for all of it. Just little kind of hacks that you could do to make your life easier… And I just let go of a lot of things. I was always a person that would do DIY on a weekend, do this, do that… I just stopped. I’ve kind of released a lot of things. It took quite a [lot of] pressure thinking I had to do this, I had to do that.”* – P1, UK

Incorporating rest and reducing energy expenditure while completing priority tasks through the use of equipment and aids was an important strategy involved in pacing. For example, sitting on a stool while cooking, or using mobility aids like canes or wheelchairs. Simple aids could be very useful, like an alarm as a reminder of a scheduled rest break.

Participants described developing acceptance and awareness of the energy required to complete an activity, and to notice patterns of symptom exacerbation to guide their pacing strategies. Some described acceptance as coming with time; others found mindfulness was a helpful strategy to cultivate awareness and acceptance. One participant described achieving a better understanding of rest through mindfulness:

*“The mindfulness… That just being in the moment and being able to kind of relax your body so your body is actually resting, because … in the beginning, I was feeling like I was resting but I wasn’t getting my energy back. But I wasn’t actually resting because I wasn’t kind of focused out. I was thinking of other things or I had the television on or something else. So yeah, I find now… that I’ve learned that, that I can actually… rest my body and I will feel rested after that. So that’s a bonus, yeah. Mindfulness is a big thing, yeah.”* – P9, Ireland

### 3) Encountering challenges with pacing

Participants described challenges with pacing in six areas including: 1) learning how to pace; 2) experiencing inequitable access to pacing; 3) experiencing stigma and judgement; 4) undergoing psychological and emotional adjustment from beliefs of ‘fighting’ or ‘pushing through’ to balancing rest with activity; 5) making sacrifices; and 6) encountering unexpected obstacles.

### Learning how to pace

Learning how to pace was difficult as pacing was a new skill for participants that they were learning while experiencing health challenges, often without formal supports. Participants described living with the health- related challenges of Long COVID for some time, experiencing ups and downs in their symptoms, before learning about pacing and being able to achieve some symptom stabilisation. Some participants felt they would have benefited from more support in the process of learning how to pace.

### Experiencing inequitable access to pacing

Another challenge was that participants experienced inequitable access to pacing as a living strategy. Some participants described feeling lucky to have the means and the freedom to plan and prioritise rest in their life to enable pacing. In addition to financial supports, access to social supports, such as daycare or help from friends/family, facilitated access to pacing. However, for participants who did not have access to such supports, planning and prioritising rest to facilitate pacing may not have been an option.

### Experiencing stigma and judgement

Many participants experienced stigma and judgement related to their efforts to pace. Participants described a lack of support from others (including friends, family, and health care providers) for pacing, stemming from poor understanding of energy limitations associated with Long COVID. Participants identified accepted cultural norms about productivity and ableism that made pacing difficult to adopt since pacing and rest are in conflict with productivity.

### Undergoing psychological and emotional adjustment from beliefs of ‘fighting’ or ‘pushing through’ to balancing rest with activity

Undergoing psychological and emotional adjustment from beliefs of ‘fighting’ or ‘pushing through’ to balancing rest with activity was challenging for many participants. Participants were torn between recognising the need for rest and the normal desire to do more in daily life.

### Making sacrifices

Participants described that pacing often involves making sacrifices. This included ‘letting go’ of previous roles, responsibilities, leisure activities, and employment, in order to pace. Sacrificing social outings and leisure activities to prioritise basic activities of daily living and essential tasks often led to participants feeling limited and isolated. Sacrifices included decisions about whether to stop working in order to effectively pace; duties involved in work might push participants outside of their energy envelope when factoring in the energy already required for basic activities of daily living and essential roles.

### Encountering unexpected obstacles

Encountering unexpected obstacles was another challenge faced when pacing. Participants’ best efforts at pacing might be derailed by unexpected life events that couldn’t be avoided or delayed. Certain events or activities were outside of their control or were essential to complete within a certain timeframe.

See Table 2 for supportive quotes for each of the challenges encountered by participants while pacing.

**Table 2:** Challenges with Pacing: Supportive Quotes.

| Challenges with Pacing | Supportive Quotes |
| --- | --- |
| Learning how to pace | *"I didn't recognise that as a cycle because to be honest I heard the term chronic fatigue syndrome but I didn't understand or know the details of it. I'd never heard the term ME. So, I didn't know anything about resting and pacing. Whenever I've had any other injury, you just exercise yourself through it right? You know, more physio and that's generally the way I think about it. So, I was sort of doing this little undulating thing throughout the fall."* – P29, Canada<br>*"Pacing is just... it is so stressful. Like I could never do like a pacing group. Like that would just not be helpful for me. I needed that one-on-one and then like it's also like hard for me to go into like the reference sheets that they give you right like that they email you like because I don't have a printer."* – P38, Canada |
| Experiencing inequitable access to pacing | *"Some people are making choices that aren't choices because... they don't have the income or the freedom... whether it be to take time off work because they just can't or just working. They shouldn't be [working], they know they shouldn't be, they've been told to rest. I think that's great; I totally agree. Also, if I don't work, I'm going to not eat, so I'm going to work. And then they have all these strategies for working to make it work but it's not working right because they're really ill and they're not going to get better at all because they're overdoing it constantly. But I get it. If you're going to choose between that and losing your home, you're going to work. I think you have to be careful about choices and a lot of people don't have them."* – P14, Canada |
| Experiencing stigma and judgement | *"I have a structure everyday... I pace myself. The physiotherapist wants me to stop sleeping in the afternoon. I disagree with her. I need that to recharge my battery, to function for the next couple of hours. She said no and I just say... I don't argue because there's no understanding of it."* – P6, Ireland<br>*"It doesn't help that my husband never sits down. He never sits down. He never rests. Yesterday his parents came up and then when I looked out the back window there was three of them up ladders cutting trees. They just, they never rest. They never sit down. He doesn't understand my need to rest really... He's trying to be understanding and all of that. But you know, 'Are you on the sofa again?' or 'Are you having another nap?' He doesn't really get it."* – P8, Ireland<br>*"We live in a very ableist world. So, it is... really hard to start to unstitch your self-worth from your productivity, to not be super hard on yourself and judge yourself as being a moral failure for not being able to clean your house, for not being able to you know cook healthy meals for your kids, for not being able to be the same active parent that you could be up until COVID."* – P25, Canada |
| Undergoing psychological and emotional adjustment from beliefs of 'fighting' or 'pushing through' to balancing rest with activity | *"For a while I was being stubborn. I was trying to fight through it and I thought I could push through this thing. But I found out to my own detriment that pushing through doesn't work. It just makes me worse, way worse..." – P10, Ireland* |
|  | *"What I ...say to people who are you know new at being long-haulers is that you have to be very patient and forgiving of yourself because there is a very human element to as soon as you start to feel remotely better, you instantly think, okay I'm cured, I can go throw myself right back into my old pre COVID routine, I am healed and boy is this place a mess from all the stuff that fell off my plate when I wasn't feeling well and I was having all these symptoms. So, I'm just going to roll up my sleeves and now that I'm feeling better again, I'm going to take care of it all in one day... you have no idea where the boundary was because you're not just inching over that line. That line is way in your rear-view mirror. You just completely fling yourself back at your regular routine... before you finally start to internalize okay my circumstances have changed, I'm not able-bodied anymore, I can't do what I used to do, I'm not back to my old baseline. And it's a huge emotional and psychological adjustment to internalize that, to acknowledge that okay maybe the day will eventually come where I'll be healthy and whole and able-bodied again. But I can't get from here to there by treating myself as able-bodied and by making the assumption that every time I'm feeling better, I can revert to all of the activities that I used to be able to get through." – P25, Canada* |
| Making sacrifices | *"...you do feel like your illness does control you. I know people do say don't let the illness control you, you control it. So, you can try and do that. You can pace and you can rest and you know you can do things bite size and with a lot of planning obviously. But the sad fact is that you know there are some things that you just cannot do. I cannot go to a restaurant... I couldn't sit upright for that long. No way. Or go to the cinema. I mean these are things that we took for granted. I haven't driven for about eight months now because I don't trust my cognition and I don't trust myself being upright. I've driven on some of those dangerous roads in the world in New Zealand and South Africa and then now you know I can't even drive the kids to school. I mean that's a huge change." – P31, UK* |
|  | *"I didn't know about pacing, which I had advice from a CBT therapist. The challenge was am I going to shower or am I going to brush my teeth today. That's how low... it's like I really changed my personality... for people who don't have chronic illness, like me, it was like I was becoming a different person. So, I'd never been in a situation where my energy was so low." – P3, UK* |
|  | *"I think the isolation... has been a huge challenge. Like you're very isolated because... you don't want to be moaning at people all the time. You know you don't want to go somewhere with someone and then say 'actually I have to sit down, you know I can't you know keep going' or you know be in a restaurant and say 'well actually I need to go home now because I'm too tired to.' I think one of the challenges is social isolation because I would have been a very social person and I would have been... obviously at work I would see lots of people and whatever and I find that a challenge." – P9, Ireland* |
|  | *"It takes all of my energy to fulfil my parental obligations when [my kids are] around and then it takes the entire week when they're gone for me to recover and have to be in the place to repeat that cycle all over again... There isn't any extra capacity for me to be working as a kindergarten teacher, even though I was only a morning only kindergarten teacher to begin with. I don't have the energy to do the physical aspect of that job, the mental aspect of that job, the emotional aspect of that job in addition to running my basics of life. I just don't have that capacity to spare yet." – P25, Canada* |
| Encountering unexpected obstacles | *"Everything has to be planned around my, I suppose, near disability you know. So, I do make a structure everyday and I try to keep it. Some days it could go off. A phone could ring and I don't answer it and then people get mad at me, 'I was ringing you all day and you never rang.' But if I take another call for half an hour to an hour on top of something else, I literally, my voice goes raspy, my oxygen drops down and I roll up like a ball in the bed." – P6, Ireland* |
|  | *"It's a lot of spoons and energy to give to managing my own healthcare... [it] should be a nice little administrative procedure but it's not. They require like all of my files and all of my information and my doctor is just not going to do it... So, I have to do it myself." – P14, Canada* |
|  | *"Sometimes it's born out of necessity, physical exertion if I have to... so there was an incident recently where there was a [parcel] that I was expecting. The courier just drove past and didn't deliver, even though I stayed home all day for it, which I normally do anyway. But then I got an email saying that it was being held at an office 2km away. I had to turn up in person and I had to bring ID... I waited for a day when I was a bit better. I took my passport. Luckily the underground station is not very far. It's about 200 metres from here. So, I took the underground and on the other side it was another 200 metres walk. I got the parcel. It was large. It was eight kilograms. I was carrying it like this. I'm obviously very cautious with money, so I didn't call a taxi. I took the subway back. Well, did I pay for it." – P31, UK* |

### 4) Experiencing consequences of not pacing

Participants described that if they did not or could not pace effectively, they faced consequences. Multiple participants referred to ‘suffering’ if they were unable to pace, or if they engaged in activities that were beyond their energy capacity. A lack of the ability or resources to pace, could lead to a relapse, crash, or exacerbation of symptoms (which could encompass physical, mental, emotional, and cognitive domains). For example, some participants would experience headaches, nausea, and cognitive challenges.

*“If I overdo it now, I would definitely have some sort of a relapse… I think that it shouldn’t be that hard to change the tire, but I suppose it is when you have something like I have… the pacing is working. Like I’ve rested now the last three weeks, but I’m still not right. I’m still not back to where I was three weeks ago… I have the fear now that I will get a relapse. I know I will if I do too much. It’s just pacing is definitely the way to go… Because if I don’t, I suffer and I mean I don’t want to suffer the way I suffered the last three weeks.”* – P10, Ireland

Symptoms experienced during a crash led to reduced function. Some participants described that they would be temporarily bed-ridden, or “*physically paralysed*” (P6, Ireland). Participants explained that the decrease in function they experienced during a crash had consequences for their ability to engage in employment and other activities. This was frustrating for participants who wanted to keep busy, or were accustomed to a higher level of activity.

*“It just feels like my brain is scrambled, like I can’t get things organised in my head anymore. I have to go lay down and not do anything which… I like being productive. I don’t like doing that. It’s like what are you supposed to do, just stop and rest? Like, I guess so. Every time I try to get out of it like sooner and my body wants to, I could feel it coming back. But the more I rest and not think about anything stressful, I think that’s akin to it feels like my mind is like it was inflamed and now it’s like calming down type thing. Once I get to a level where it’s low enough in terms of like this so-called inflammation in my mind, I’m able to work again. It takes about two to three days.”* – P23, US

There was also tangible pressure experienced to continue to engage in employment, as being unable to do so resulted in lost income and financial insecurity.

*“I lost like $10,000 of my salary because of choices I had to make because I had to manage Long COVID… So, there’s a pressure to maintain wellness… because I can’t afford to not be well. So that can be a bit of a pressure but it also helps to have a bit more discipline and pace in myself to try to stay well.”* – P1, UK

### 5) Conceptualising and describing pacing using analogies and metaphors

Participants used a range of analogies and metaphors to conceptualise and to describe the process of pacing. Examples of analogies and metaphors used by participants included Spoon Theory, budgeting energy, staying within an ‘energy envelope’ and ‘stoplight’ (see Figure 2). Some participants articulated the importance of using analogies and metaphors to describe pacing to others. This helped to validate the utility of pacing and to make it more tangible (for themselves and others). For example, *“[Spoon Theory is] a really helpful way of just naming what you’re working with.”* (P40, US).

**Figure 2:**
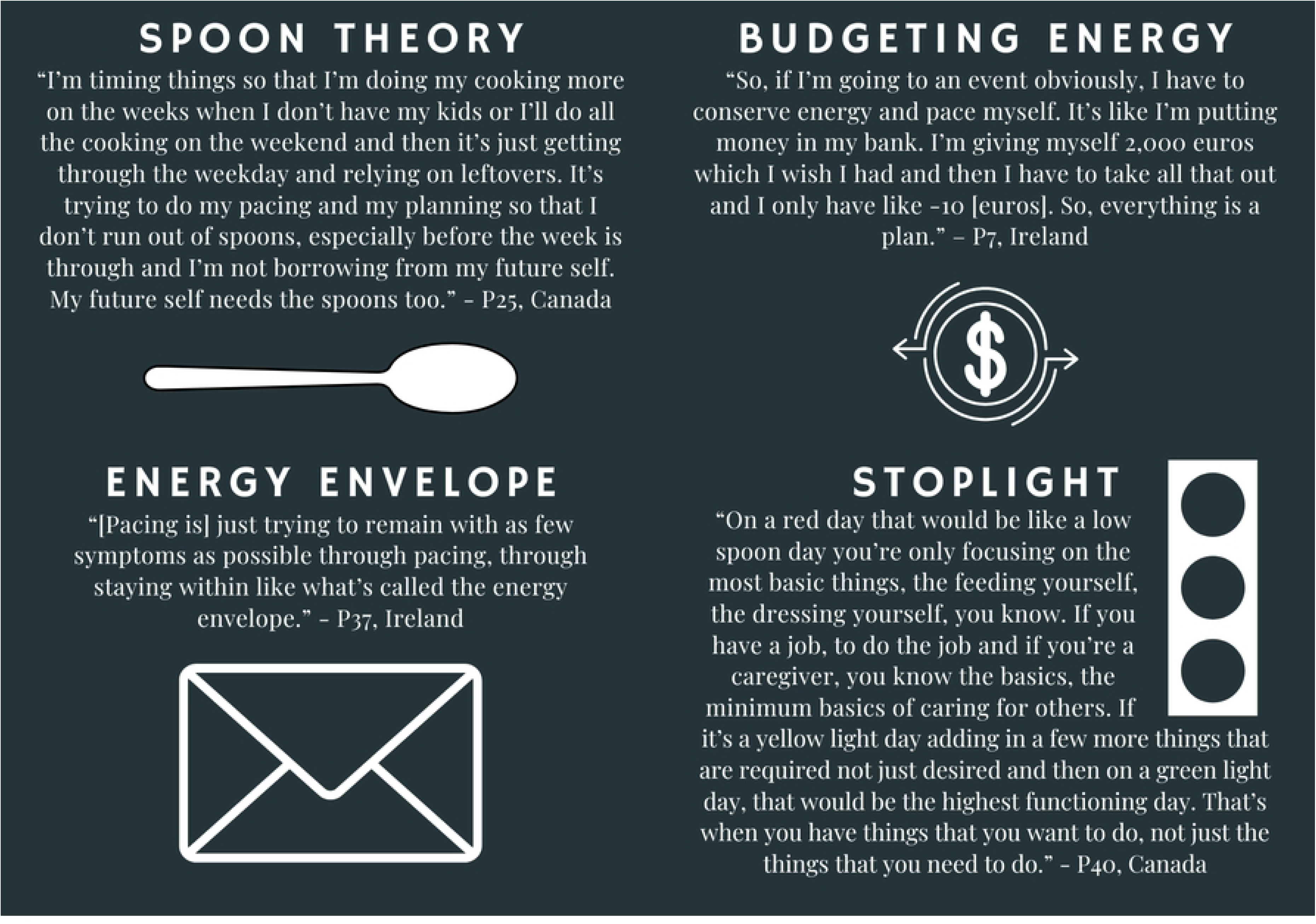
Analogies and Metaphors for Pacing

### Spoon Theory

Spoon Theory is based on the premise that only a finite number of spoons can be allocated to activities throughout the day, and day-to-day the number of spoons available can change (41). Participants who knew about and discussed Spoon Theory recognised its origins from within the community of people living with chronic disability (Figure 2). As this participant eloquently describes, Spoon Theory is a helpful analogy to describe pacing in the context of episodic disability.

*“Spoon Theory, I’m going to [forget] the name of the person who came up with it… Anyway, it was someone with chronic illness who came up with spoon theory as a way of explaining the need to do pacing when it comes to how much energy you have to give and that it isn’t a solid block of energy that will be consistent every single day. It’s more like episodic illness where some days you have say a bank of 10 spoons to use to get you through your activity and different activities will require more spoons or fewer spoons. Say eating is one spoon and driving is three spoons, a conversation on the phone is three spoons if it’s social but if it’s the bank for insurance it might be five and so on and so forth. So… as you get to be more familiar and more aware with what your energy capacity is and how much any given activity takes out of you, you get better over time with planning how much energy to use on any given day.”* – P25, Canada

Participants communicated important insights about pacing using Spoon Theory. For example, one participant used Spoon Theory to communicate the sacrifices they made about what roles they could take on in life, based on their limited number of spoons:

*“Could I work if I wasn’t a single mom? Probably not. But it’s just I only have like say 20 spoons a day. The rest is 100. I might have 20 on the most amazing day. Usually, it’s like 10. So, I’m spending eight of those on my kid and like life activities. There’s just not much left for anything else at the moment and I don’t have a lot of support too.”* – P14, Canada

### Budgeting energy

Several participants compared pacing and budgeting, “*knowing like how much energy I have, what I want to spend it on” (P40, US),* to not end up in an energy deficit or debt. Participants described the process of ‘saving their energy’ through rest, and ‘spending their energy’ on tasks, emotion or physical exertion (Figure 2).

*“If you use a lot of emotional energy, there’s a lot of stuff going on around you, just your emotions or whatever, that can take a lot from you as well. [There are] three prongs to that. But definitely I have been better at knowing how to save my energy. It’s not like I can just like say okay I rested yesterday, so I’ll be fine today. That’s not the way it is… Whereas if I used up a lot of energy yesterday, if I did… like if I went shopping yesterday, you know I might not be as able to do this [interview] today.”* – P9, Ireland

### Energy Envelope

Some participants in this study described pacing in order to remain within the ‘energy envelope.’ The envelope represented the amount of energy available to someone. Expending more energy than available will exceed the limits of the envelope (Figure 2).

### Stoplight

One participant described a stoplight system that they used to guide their pacing efforts. They described identifying each given day as a ‘red’, ‘yellow’ or ‘green’ day. Red days represented low energy days; on these days, they would only perform basic necessary activities of daily living to avoid triggering symptom exacerbation. Green days represented higher energy days; on these days, they could engage in higher level functional activities and leisure activities. Yellow days would fall in between (Figure 2).

### Other analogies and metaphors

Several other analogies and metaphors were used by participants to conceptualise and describe pacing. For example, participants described having a *“body battery”* (P40, US) that is drained by exertion and charged by rest. One participant described *“living in a land of eggshells”* (P4, UK) where “*I feel like I’ve got to hold myself back a little bit, like I’ve got to just slowly step on eggshells”* (P4, UK). This metaphor of walking on eggshells communicated the restraint required for pacing.

## DISCUSSION

This study set out to describe the experiences of pacing among adults living with Long COVID in Canada, Ireland, UK, and US. Participants experienced pacing as being a helpful living strategy to prevent or reduce symptom exacerbation. Many were knowledgeable, and had learned about pacing through online community support groups for people living with Long COVID or other energy-limiting chronic conditions. However, the experiences of pacing were described as a ‘moving target’, involving many challenges, and requiring time, effort, and trial and error to implement effectively. Participants used analogies and metaphors to conceptualise and describe pacing, such as Spoon Theory, budgeting their energy, and staying within their energy envelope.

This study builds on previous work demonstrating that pacing is often a new concept for people living with Long COVID, and that learning to pace is challenging but critical in symptom management (42, 43). Participants in this study, described pacing as requiring extensive trial and error to find and maintain a balance between rest and exertion, adapting to fluctuations in energy levels and the presence and severity of symptoms over time. Participants also described that experiencing social inequities, and stigma or judgment posed substantial challenges to pacing. This points to the need for accessible social and disability supports to reduce barriers and challenges to implementing pacing (44, 45).

Findings from this study highlight how community support groups have been instrumental to participants’ experiences of learning how to pace, specifically by acquiring knowledge about pacing. Support groups are helping individuals living with Long COVID to learn about pacing and to navigate challenges faced with pacing, while many health and rehabilitation providers remain uninformed about pacing resulting in scarcity of formalised supports for pacing (23). Support and community groups are instrumental in helping people living with Long COVID to learn about and implement pacing (23, 46). People living with Long COVID came together through online platforms and support groups early in the pandemic, before academic and medical attention had moved beyond the acute stages of COVID-19 (47). These platforms have remained valuable for people living with Long COVID; they have driven advocacy efforts and provided educational resources and social and emotional support (46, 48–50). However, some individuals living with Long COVID, particularly those experiencing cognitive impairment, may struggle to navigate the substantial amount information available on support groups, which may trigger feelings of overwhelm (46). Additionally, the onus to maintain and coordinate support groups requires effort and energy on the part of those with lived experiences who are leading the groups (23, 46). It may be particularly challenging for the organizers to attempt to field complex questions about pacing and other living strategies, and to provide individualized information. Despite potential limitations, peer-support groups for Long COVID remain a tremendous and valued resource, particularly as health care providers continue to build capacity and knowledge to more adequately meet the needs of people living with Long COVID (46, 51).

Participants described experiencing difficulty accessing health and rehabilitation providers who were knowledgeable about pacing in a timely manner. There is a need to educate health and rehabilitation providers about pacing as a supported self-management or living strategy for Long COVID, to increase access to specialised providers and clinics, and to facilitate access to community support groups (51). Evidence suggests that Long COVID support groups are disproportionally utilized by white people aged 35-55 years (46), resulting in many people living with Long COVID with limited access to information shared in support groups. Participants in this study described that health and rehabilitation professionals are lacking in knowledge about pacing, despite existing guidelines and online resources (29, 50); participants were left waiting for lengthy periods of time to access specialised providers or clinics who could provide tailored guidance on managing Long COVID, echoing previous findings (52). Enhancing education of health and rehabilitation providers on Long COVID and living strategies, such as pacing, could help people living with Long COVID manage their symptoms better and feel more supported in their communities (53). Investing in and expanding access to specialised Long COVID clinics and providers also is important to ensure that people living with Long COVID are able to access more in-depth and tailored support for pacing as needed (52).

Given that support groups will continue to fill gaps in knowledge about Long COVID management, including pacing, it is essential that they are engaged in research and knowledge translation efforts related to pacing, and that resources are directed toward these groups to support them in delivering critical information on pacing and other living strategies for people living with Long COVID (46).

Results in this study also highlight that the opportunity and ability to implement pacing and prioritise rest as a meaningful activity, are not equal for everyone. To prioritise rest, participants described needing to take time off work or to depend on family, friends, or caregivers to assist them with essential tasks and roles, such as housekeeping and childcare. This is consistent with the results of other research on the impact of Long COVID on employment and pacing (54, 55). If participants were unable to access employment or disability benefits, they were faced with the impossible choice between continuing to struggle with their health versus sacrificing financial stability to take time off work. Not all participants possessed strong social networks to help with childcare and day-to-day activities. Social inequities may influence the ability to prioritise and balance rest with activity, and impact access to healthcare, information and support for living strategies for people with Long COVID, as with other chronic illnesses (56–58). Therefore, some adults living with Long COVID may face greater challenges learning about pacing and accessing support to implement pacing compared to others. It is vital that work accommodations, insurance and disability benefits, and home and community-based services are made accessible to people living with Long COVID to facilitate implementation of pacing as a living strategy to reduce symptom exacerbation (44, 59–61).

The analogies and metaphors used by participants in this study to conceptualise pacing and to describe it to others may be powerful communication tools for other people living with Long COVID and other energy-limiting conditions. People living with Long COVID have broadly reported dismissal and invalidation of their symptoms by healthcare providers, friends, and family members (62–65). Validation of symptoms is an important aspect of care that people living with Long COVID are seeking from health providers (66–68). Rest, a meaningful activity required for pacing, is undervalued in the context of contemporary ‘hustle culture’, which prioritises productivity and busyness (69). Therefore, efforts to pace may be dismissed, minimized or criticized by others. This was the case for many of the participants in this study; even participants themselves had to undergo psychological adjustment to embrace pacing. For people living with Long COVID, their healthcare providers, and their communities to understand pacing, it is important to have clear and relatable means of communicating what pacing is and how it works. Analogies and metaphors shared by participants in this study, such as balancing activities and rest, Spoon Theory, budgeting energy, and staying within the energy envelope, are examples of how information about pacing can be effectively communicated with providers and caregivers.

Strengths of this study include the inclusion of participants living with Long COVID in four countries, our community-engaged team-based approach, and community-led recruitment with purposive sampling to achieve diversity in gender, sexual orientation, and race. The aim of the parent study was to describe experiences of disability among adults living with Long COVID, and not specifically to explore experiences with pacing (11). However, the amount and the richness of data on the experiences of pacing from 37 out of 40 of the participants highlighted the importance and utility of this living strategy. While participants were from Canada, Ireland, UK, and US, our aim was to not compare and contrast between countries; instead we analyzed interview data as one collective dataset. Recruitment through community support groups fostered trust among participants and likely contributed to the richness of interview data. However, as participants were actively engaged in community support groups and networks, they may have been more knowledgeable about Long COVID, pacing, and existing supports than the broader population of persons living with Long COVID. Finally, our study was focused in four high-income settings; results may not be transferable to low- and middle-income settings, but provide important foundations to further explore pacing in other contexts.

Future directions for research on pacing should include studies that explore the potential for technology to facilitate pacing for adults living with Long COVID, and studies that assess the co-creation and implementation of pacing strategies and programming into healthcare systems and social support institutions. People living with Long COVID should be engaged in the design and implementation of future research on pacing (68, 70). People living with Long COVID have used smart watches and other technology to guide their pacing efforts (71). Research on pacing as a rehabilitation strategy and intervention and the role of technology to support pacing may help to develop evidence-based guidelines that can be implemented by people living with Long COVID and their health care providers (72). Research should also explore knowledge, experiences, and utility of pacing among populations who are under-represented in this study and other research on pacing, such as adolescents, children, and young people living with Long COVID, people living in low- and middle-income countries, and people from underrepresented racial and sexual and gender groups.

## CONCLUSION

Pacing is a critical strategy used by people living with Long COVID to mitigate, manage and stabilize symptoms; however, pacing is experienced as a “moving target” for adults living with Long COVID. Even after learning about pacing, adults living with Long COVID may experiences challenges with pacing as well as consequences of not pacing. Adults living with Long COVID use analogies and metaphors to conceptualise and describe pacing, for enhancing understanding and support for pacing. Health and rehabilitation providers should consider that pacing is challenging and complex, and work collaboratively with patients to implement this strategy, when appropriate.

## Data Availability

All relevant data are within the manuscript and its Supporting Information files.

## ACKNOWLEDGEMENTS

We thank the participants for their contributions to this study and the community organizations who collaborated in this work. We acknowledge the following community collaborator organizations who were instrumental in this study: the Patient-Led Research Collaborative (PLRC), Long COVID Physio, Long COVID Support UK, COVID Long Haulers Support Group Canada, Long COVID Advocacy Ireland, and Long COVID Ireland.

## Notes

### Competing Interest Statement

The authors have declared no competing interest.

### Author Declarations

The study was approved by the University of Toronto Health Sciences Research Ethics Board (protocol #41749).

